# Time to death and its Predictors among Adult TB/HIV Co-Infected Patients in Mizan Tepi University Teaching Hospital, South West Ethiopia

**DOI:** 10.1101/19004234

**Authors:** Wondimagegn Wondimu, Lamessa Dube, Teshome Kabeta

## Abstract

**Background:** Tuberculosis (TB) and Human Immuno Deficiency Virus (HIV) co-infection represents a complex pathogenic scenario with synergistic effect and leads to about 300,000 HIV-associated TB deaths in the world in 2017. Despite this burden of death, time to death and its predictors among TB-HIV co-infected patient was not adequately studied; and the existing evidences are inconsistent. Therefore, this study was aimed to determine time to death and identify its predictors among adult TB/HIV co-infected patients.

**Method:** Retrospective cohort study was conducted by reviewing registers of randomly selected 364 TB/HIV co-infected patients enrolled in health care from July 2, 2007 up to July 1, 2017 at Mizan Tepi University Teaching Hospital. The hospital was located in Bench Maji Zone, South West Ethiopia. Data were collected from March 1 through 31, 2018, entered to Epi data 3.1 and exported to SPSS version 21. Each patient was followed from date of TB treatment initiation till death, loss to follow up and treatment completed. On the other hand, events other than death were considered as censored. After checking the proportional hazard model assumption, Cox-regression was used to identify the predictors. In bivariable analyses, P-value≤0.25 was used to identify candidate variables for multivariable analysis. The 95% CI of hazard ratio (HR) with respective P-value <0.05 was used to declare significance in the final model.

**Result:** All the 364 patients were followed for 1,654 person months. There were 83 (22.8%) deaths and most 38 (45.8%) were occurring within the first two months of anti-TB treatment initiation. The overall incidence rate and median survival time were 5.02 per 100 person months (95% CI: 4.05, 6.22) and 10 months respectively. Statistically significant better survival was observed among patients: with CD4 ≥ 200 cells/mm^3^ (P<0.001), who had a history of cotrimoxazole preventive therapy (CPT) use (P<0.001), who disclose their HIV status (P<0.001) and with working functional status (P<0.001). Not using CPT (adjusted hazard ratio [AHR] =1.72; P=0.023), bedridden functional status (AHR=2.55; P=0.007), not disclosing HIV status (AHR=4.03; P<0.001) and CD4 < 200 cells/mm^3^ (AHR=6.05; P<0.001) were predictors of time to death among TB/HIV co-infected patients.

**Conclusion:** The median survival time was 10 months and poor survival was associated with low CD4 count, not using CPT, not disclosing HIV status and having bedridden functional status. Close monitoring of bedridden and low CD4 count patients, prompt CPT initiation and encouraging HIV status disclosure are recommended.

## Introduction

TB/HIV co-infection has dangerous synergy which leads to highly complicated features of both diseases, including epidemiologic profile, pathogenesis, clinical presentation, treatment, and prevention methods (1). It has also social, economic and political crisis as consequences. The most common consequences are stigma, high susceptibility to other comorbidities like mental disorders, cost of care, increased number of orphans and decreased productivity due to long time illness and absence from work place (2,3). In most of the situations, this co-infection affects the productive segment of the population and increases the crisis by many folds, especially for low income countries like Ethiopia (4).

For people living with HIV/AIDS (PLWHA), the risk of developing TB is estimated to be between 17-23 times greater than those without HIV infection in 2017. Globally, the proportion of HIV-positive patients who died during TB treatment was 11%, in the same year (5,6).

TB was the leading cause of death among people with HIV and there were about 300,000 people who died from HIV-associated TB in 2017 in which the majorities of deaths (84%) occurred in Africa (6).

Ethiopia is among the 30 high TB/HIV burden countries and among TB patients with known HIV status, which is about 11%. It was estimated that there were about 3600 deaths due to TB-HIV co-infection in 2017 (5,7). On the top of knowledge gap, the problems of lay beliefs, misconceptions and prejudices regarding TB/HIV co-infection are other devastating issues in Ethiopia; since this all might contribute to delay from care and in turn for poor survival among co-infected patients (8–10).

Currently, there is no published study in the study area on time to death and its predictors among TB/HIV co-infected patients; although it is immediate near to Gambella region, which is among the regions with higher TB/HIV co-infection in Ethiopia and from where also a significant number of TB/HIV patients come to Mizan-Tepi University Teaching Hospital (MTUTH).

In addition, studies conducted in North West and South West part of Ethiopia identified inconsistent predictors (11,12). This study was aimed to determine time to death and its predictors among TB-HIV co-infected adult patients in MTUTH.

## Methods

### Study Design and Setting

A retrospective cohort study was conducted among TB/HIV co-infected patients enrolled from July 2, 2007 to July 1, 2017 at Mizan Tepi University Teaching Hospital. Each patient was followed from date of TB treatment initiation till death, loss to follow up and treatment completed. On the other hand, events other than death were considered as censored.

The hospital is in Mizan-Aman town, which is 593 km to South West of Addis Ababa, capital of Ethiopia. The hospital started offering free ART service in 2006. About 5946 PLWHA were enrolled till January 2019 and among these 1830 were active. Assessment for TB was being done routinely for every HIV patient. Similarly, if their test results become positive, all TB patients were supported to know their HIV status and linked to ART clinic.

### Study Population and Sampling Techniques

All adult TB/HIV co-infected patients, who received care from MTUTH from July 2, 2007 to July 1, 2017 were included. Incomplete registries, transfer-out and death in the same day of anti-TB treatment initiation were excluded.

The sample size was determined using the online calculator for test time-to-event data for the Cox proportional hazard (13). The CD4 count ≤50 is known predictor of mortality and used to classify the groups into two (i.e. subjects with CD4 count ≤50 cells/mm^3^ and subjects with CD4 count >50 cells/mm^3^) (4,14). Using the parameters (power=80%, type one error rate=5%, hazard ratio=2.31, overall probability of event=13% and proportion of sample in group of subjects with CD4 count ≤50 cells/mm^3^=38.7%) from previous study (15), the sample size became 364. Final samples were selected by simple random sampling technique after listing medical registration number of all 670 eligible records as a sampling frame.

### Data Collection Technique and Data Quality Control

Data were collected using a data extraction tool adapted from national ART and anti TB treatment standard registries. Data sources were standard registries of ART and anti-TB treatments, electronic format, patient medical record (card) and intake forms. Two trained nurses each from the ART and TB clinics of the hospital collected the data. Trained supervisor supervises all the collection process. Both the supervisor and principal investigator daily checked the completeness and consistency of checklists during data collection. Socio demographic, clinical, behavioural and health service-related base line data were collected from the sources.

### Data entry, processing and analysis

Data were entered to Epi data 3.1 and exported to SPSS version 21 for analysis. Exploration of the data was done to check the missing values, outliers, and proportionality of hazards over time. Continuous variables were described using means and medians with respective 95% CI; while categorical variables were described using counts, percentages and tables. The median survival time, survival probability and the incidence rate were determined. The association between the independent variables and outcome variable was assessed by Cox-proportional hazard model and hazard ratio (HR) was used as a measure of association. Survival status of TB/HIV co-infected patients was presented by life table.

All variables with p-value ≤0.25 were candidates for the final model. By using backward stepwise likely hood ratio technique and p-value <0.05 the final significant predictors were identified.

### Ethical consideration

The study proposal was approved by an institutional review board (IRB) of Jimma University. Support letter was obtained from IRB of Jimma University to MTUTH administrator’s office. Informed consent was obtained from the concerning administrative body of the hospital. Confidentiality was assured strictly. In addition, patient identifiers (medical registration number and ART unique number) were replaced by a new identification number during data collection and the name of the patients was not used.

## Results

### Characteristics of cohort

Among eligible 670 patients, 364 were included to the study. The mean age (±SD) of the participants was 30.52(±9.03). From the total participants, 211(58%) were females, 150(41.2%) were married and 133(36.5%) had attended secondary school. **(Table 1**)

**Table 1.** Base line socio demographic characteristics of study subjects in MTUTH, South West Ethiopia, 2007-2017

| Variable | Variable category | Frequency | Percent |
| --- | --- | --- | --- |
| Age | 15-24 | 91 | 25 |
|  | 25-34 | 160 | 44 |
|  | 35-44 | 81 | 22.2 |
| | ≥45 | 32 | 8.8 |
| Sex | Male | 153 | 42 |
|  | Female | 211 | 58 |
| Educational status | No education | 70 | 19.2 |
|  | Primary (grade 1-8) | 80 | 22 |
|  | Secondary (grade 9-12) | 133 | 36.5 |
|  | Tertiary (college/university) | 81 | 22.3 |
| Marital status | Never married | 140 | 38.4 |
|  | Married | 150 | 41.2 |
|  | Divorced | 37 | 10.2 |
|  | Widowed | 37 | 10.2 |
| Occupation | Gov't employed | 48 | 13.2 |
|  | Private organization employed | 31 | 8.5 |
|  | Jobless | 44 | 12.1 |
|  | Student | 46 | 12.6 |
|  | Farmer | 35 | 9.6 |
|  | Commercial sex worker | 42 | 11.5 |
|  | Merchant | 83 | 22.8 |
|  | Housewife | 35 | 9.6 |
| **Place of residence** | Within catchment area | 261 | 71.7 |
|  | Out of catchment area | 103 | 28.3 |
| **Having HIV-positive** | No | 100 | 27.6 |
| **family member (n=362) \*** | Yes | 262 | 72.4 |

\* Two participants had no records on either they had HIV positive family member or not.

Participants with baseline CD4 count at least 200 cells/mm were 240 (65.9%). Significant portion of study participants had one or two opportunistic infections other than TB, 232(63.7%). Cotrimoxazole was used as preventive therapy among 283 (77.7%) of the participants. **(Table 2**)

**Table 2.**
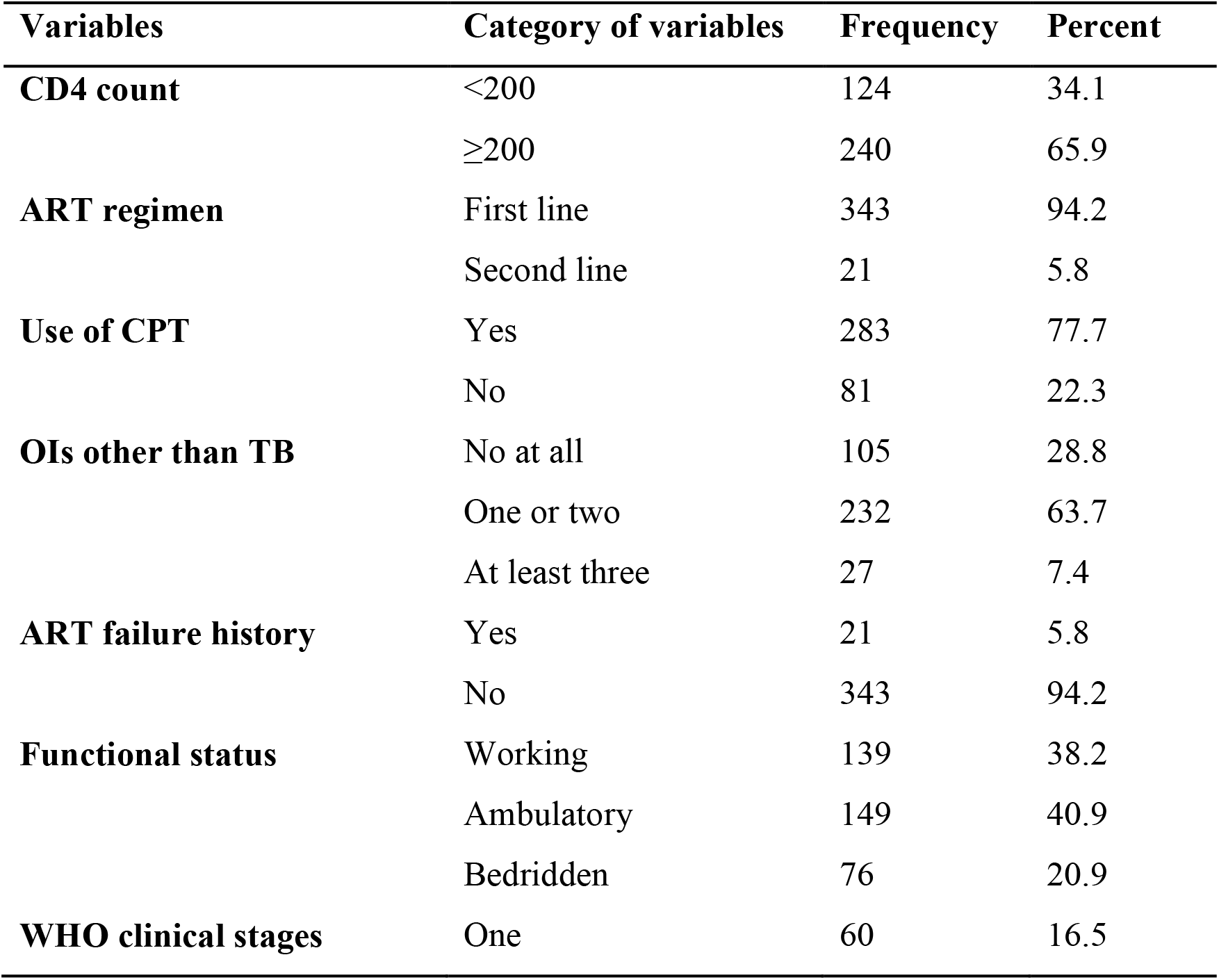
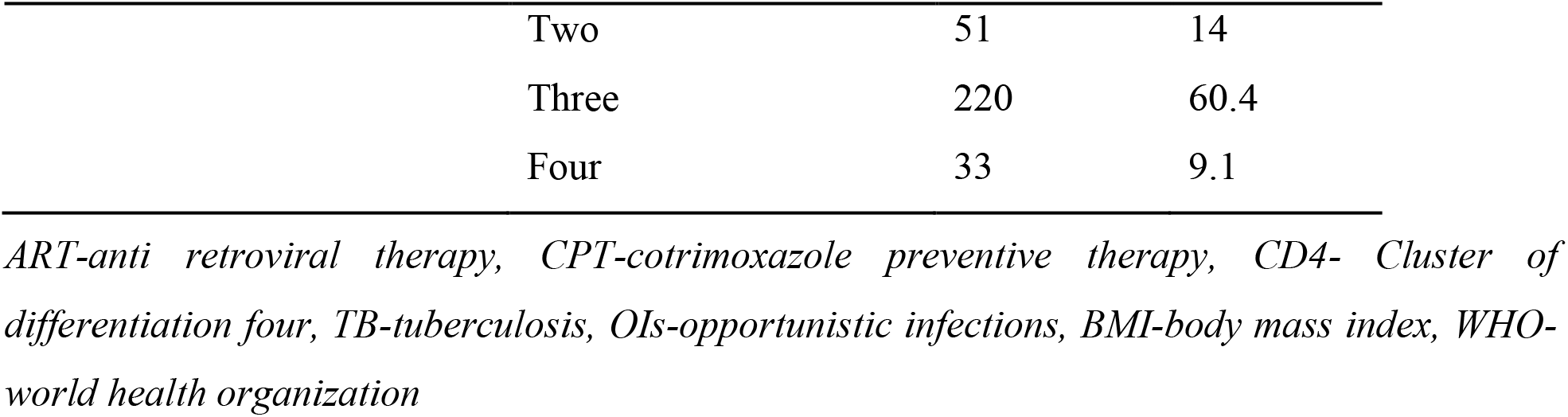
Clinical characteristics of study participants in MTUTH, South West Ethiopia, 2007-2017

| **Variables** | **Category of variables** | **Frequency** | **Percent** |
| --- | --- | --- | --- |
| **CD4 count** | <200 | 124 | 34.1 |
|  | ≥200 | 240 | 65.9 |
| **ART regimen** | First line | 343 | 94.2 |
|  | Second line | 21 | 5.8 |
| **Use of CPT** | Yes | 283 | 77.7 |
|  | No | 81 | 22.3 |
| **OIs other than TB** | No at all | 105 | 28.8 |
|  | One or two | 232 | 63.7 |
|  | At least three | 27 | 7.4 |
| **ART failure history** | Yes | 21 | 5.8 |
|  | No | 343 | 94.2 |
| **Functional status** | Working | 139 | 38.2 |
|  | Ambulatory | 149 | 40.9 |
|  | Bedridden | 76 | 20.9 |
| **WHO clinical stages** | One | 60 | 16.5 |

|  |  |  |
| --- | --- | --- |
| Two | 51 | 14 |
| Three | 220 | 60.4 |
| Four | 33 | 9.1 |

*ART-anti retroviral therapy, CPT-cotrimoxazole preventive therapy, CD4- Cluster of differentiation four, TB-tuberculosis, OIs-opportunistic infections, BMI-body mass index, WHO-world health organization*

### Survival status and its predictors

Three hundred sixty-four participants were followed for total of 1,654 person months. There were 83 (22.8%) deaths and 41(11.3%) loss to follow up. From the total deaths, 38 (45.8%) occurred within the first two months of anti-TB treatment. The overall incidence rate of death was 5.02 per 100 person months (95% CI: 4.05, 6.22). The median survival time was at around the 10^th^ month. Survival at 2, 6 and 12 months after initiating TB treatment were 82.02%, 73% and 45%respectively. **(Table 3)**

**Table 3.**
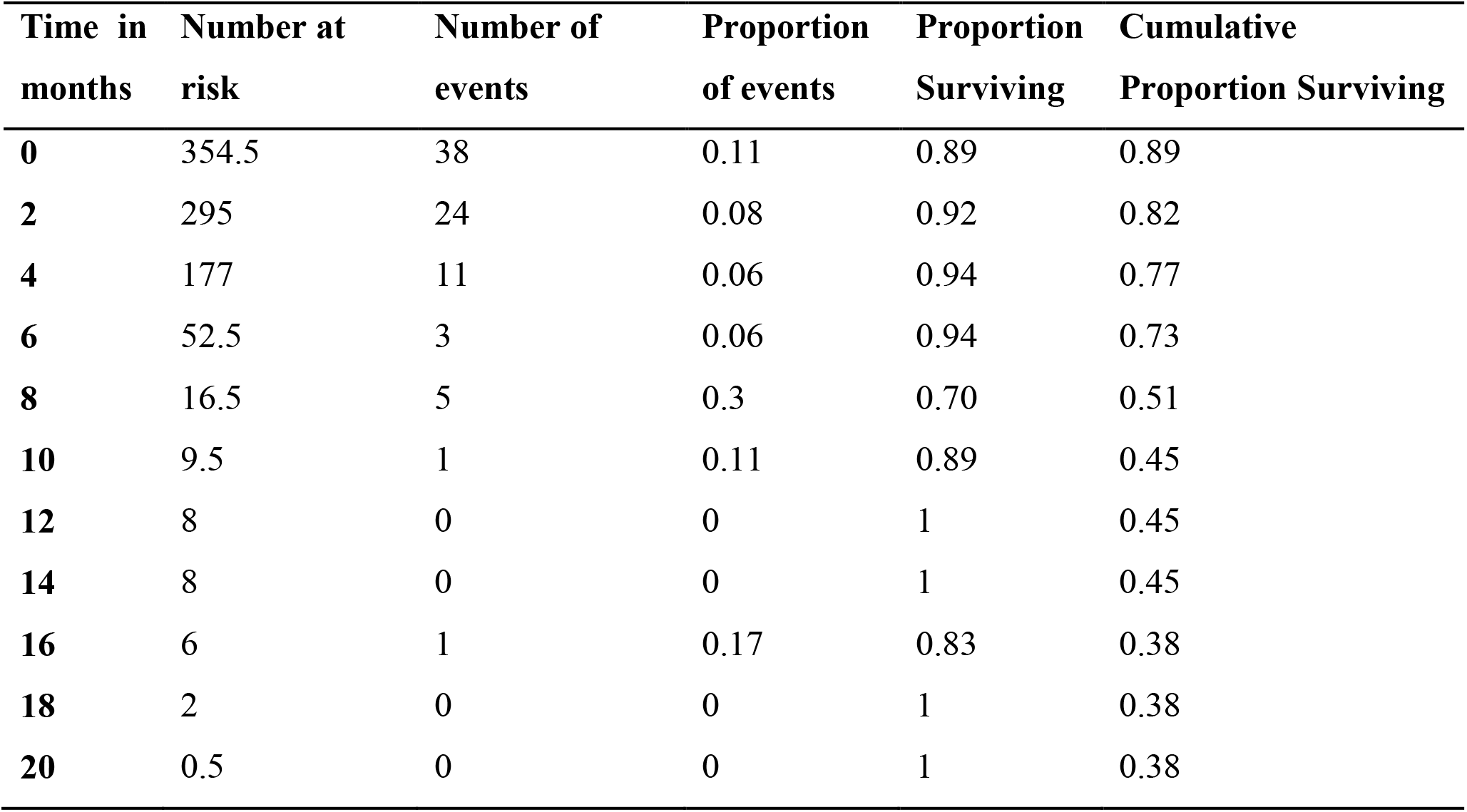
Overall life table of TB/HIV co-infected patients in MTUTH, South West Ethiopia, 2007-2017

The significant predictors of time to death for TB/HIV co-infected patients were; not using CPT (AHR=1.72; 95% CI: 1.08, 2.74), bedridden functional status (AHR=2.55; 95% CI: 1.29, 5.06), not disclosing HIV status (AHR=4.03; 95% CI: 2.21, 7.35) and CD4 count of less than 200 cells/mm^3^. (**Table 4**)

**Table 4.**
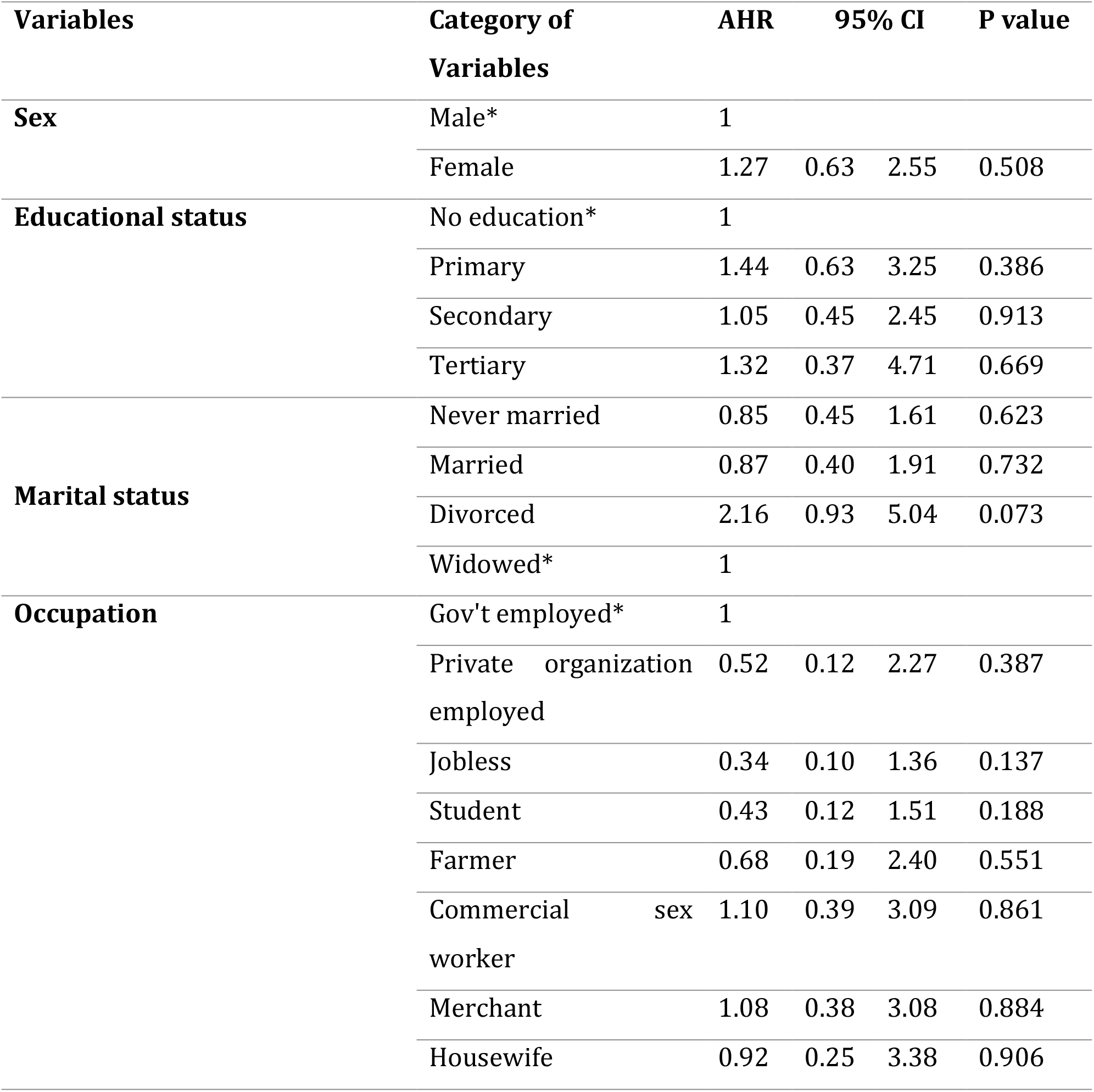
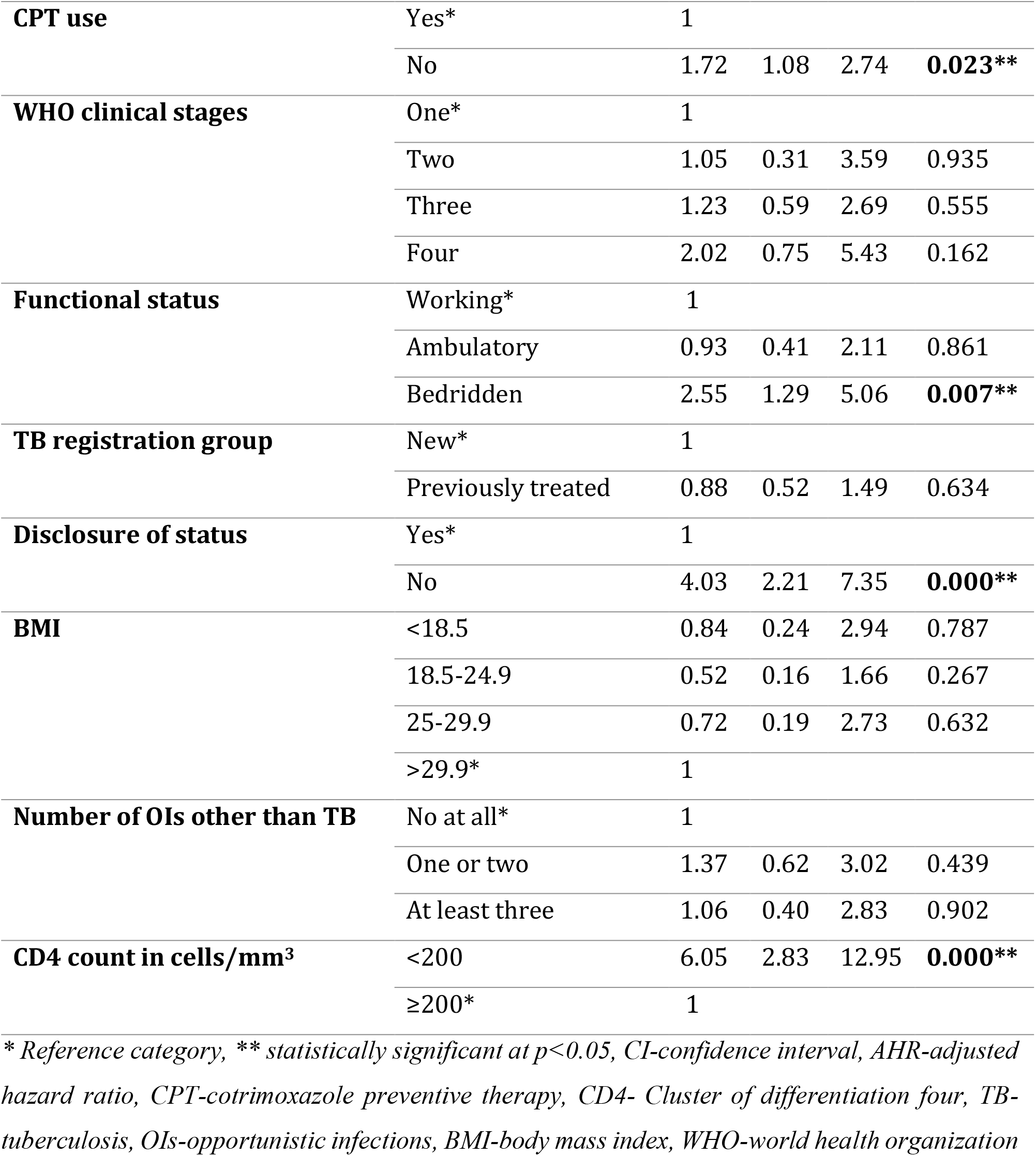
Multivariable cox- regression analysis for predictors of time to death among TB/HIV co-infected patients in MTUTH, South West Ethiopia, 2007-2017

| Variables | Category of Variables | AHR | 95% CI |  | P value |
| --- | --- | --- | --- | --- | --- |
| **Sex** | Male* | 1 |  |  |  |
|  | Female | 1.27 | 0.63 | 2.55 | 0.508 |
| **Educational status** | No education* | 1 |  |  |  |
|  | Primary | 1.44 | 0.63 | 3.25 | 0.386 |
|  | Secondary | 1.05 | 0.45 | 2.45 | 0.913 |
|  | Tertiary | 1.32 | 0.37 | 4.71 | 0.669 |
| **Marital status** | Never married | 0.85 | 0.45 | 1.61 | 0.623 |
|  | Married | 0.87 | 0.40 | 1.91 | 0.732 |
|  | Divorced | 2.16 | 0.93 | 5.04 | 0.073 |
|  | Widowed* | 1 |  |  |  |
| **Occupation** | Gov't employed* | 1 |  |  |  |
|  | Private organization employed | 0.52 | 0.12 | 2.27 | 0.387 |
|  | Jobless | 0.34 | 0.10 | 1.36 | 0.137 |
|  | Student | 0.43 | 0.12 | 1.51 | 0.188 |
|  | Farmer | 0.68 | 0.19 | 2.40 | 0.551 |
|  | Commercial sex worker | 1.10 | 0.39 | 3.09 | 0.861 |
|  | Merchant | 1.08 | 0.38 | 3.08 | 0.884 |
|  | Housewife | 0.92 | 0.25 | 3.38 | 0.906 |
| **CPT use** | Yes* | 1 |  |  |  |
|  | No | 1.72 | 1.08 | 2.74 | **0.023\*\*** |
| **WHO clinical stages** | One* | 1 |  |  |  |
|  | Two | 1.05 | 0.31 | 3.59 | 0.935 |
|  | Three | 1.23 | 0.59 | 2.69 | 0.555 |
|  | Four | 2.02 | 0.75 | 5.43 | 0.162 |
| **Functional status** | Working* | 1 |  |  |  |
|  | Ambulatory | 0.93 | 0.41 | 2.11 | 0.861 |
|  | Bedridden | 2.55 | 1.29 | 5.06 | **0.007\*\*** |
| **TB registration group** | New* | 1 |  |  |  |
|  | Previously treated | 0.88 | 0.52 | 1.49 | 0.634 |
| **Disclosure of status** | Yes* | 1 |  |  |  |
|  | No | 4.03 | 2.21 | 7.35 | **0.000\*\*** |
| **BMI** | <18.5 | 0.84 | 0.24 | 2.94 | 0.787 |
|  | 18.5-24.9 | 0.52 | 0.16 | 1.66 | 0.267 |
|  | 25-29.9 | 0.72 | 0.19 | 2.73 | 0.632 |
|  | >29.9* | 1 |  |  |  |
| **Number of OIs other than TB** | No at all* | 1 |  |  |  |
|  | One or two | 1.37 | 0.62 | 3.02 | 0.439 |
|  | At least three | 1.06 | 0.40 | 2.83 | 0.902 |
| **CD4 count in cells/mm<sup>3</sup>** | <200 | 6.05 | 2.83 | 12.95 | **0.000\*\*** |
|  | ≥200* | 1 |  |  |  |

\* Reference category, \*\* statistically significant at $p < 0.05$ , CI-confidence interval, AHR-adjusted hazard ratio, CPT-cotrimoxazole preventive therapy, CD4- Cluster of differentiation four, TB-tuberculosis, OIs-opportunistic infections, BMI-body mass index, WHO-world health organization

## Discussion

This study revealed that there were 22.8% deaths among TB/HIV co-infected patients during TB treatment and this finding is comparable to the studies done in Jimma and Bahirdar towns where there were 20.2% and 22% deaths respectively. The overall incidence rate was 5.02 per 100 person months, which is comparable to a study done in Bahirdar in which there was 4.09 per 100 person months incidence rate during TB treatment (12,16). The comparability of death rates in these all studies is suggestive for Ethiopia’s rank of being among TB/HIV high burden countries in the world and the need of future high attention and priority particularly for the timely achievement of ending both epidemics (7,17).

The majority of the deaths, 38 (45.8%) occurred within the two months of anti-TB treatment initiation and this is fairly comparable with a study conducted in Malaysia where 40% of deaths occurred in the same period of TB treatment (18). This is suggestive of the need of appropriate management of drug interactions and overlapping toxicities of anti TB and ART drugs, immune reconstitution inflammatory syndrome (IRIS) and other OIs which were factors highly associated with early deaths in TB/HIV co-infected patients (19–21).

The survival at 2, 6 and 12 months after initiating TB treatment were 82.02%, 73% and 45% respectively. This is lower than the Malaysian finding where there were survival of 90.7%, 82.8% and 78.8% respectively at 2, 6 and 12 months after initiating TB treatment (18). This might be due to the pioneering efforts in Malaysia to combat TB/HIV co-infection; which is evidenced by its early achievement of the first and third 90s of UNAIDS’ 90-90-90 target (22).

Patients who didn’t use CPT were nearly two times (AHR=1.72) at higher risk of death and in agreement with this, study conducted in Bahirdar revealed that not taking CPT was significantly associated with mortality among TB/HIV co-infected patients during TB treatment (12). Severe bacterial infections and malaria, which can be successfully prevented by CPT might be attributable to higher risk of death among CPT non users (14,23,24).

Bedridden patients were two and a half times (AHR=2.55) risky to die than those who had working functional status. This is similar to findings of studies conducted in Jimma and Ambo (11,25,26). This might be due to the delayed presentation of bedridden patients to care after developing numerous infections and complications which were in turn associated with poor survival (10,18).

The bidirectional and synergistic effect of TB and HIV infection has profound impact on the survival of TB/HIV co-infected patients. Latent or active TB activates CD4 cells to be captured by HIV and HIV depletes CD4 cells, which in turn accelerates the progression of latent TB to active TB disease (1,27). CD4 count of less than two hundred cells per cubic millimeter was significantly associated with poor survival of TB/HIV co-infected patients in this study (AHR=6.05). This is supported by previous findings from different parts of the world including Ethiopia. Although the current estimation is consistent with the findings of Bahirdar and Malaysia, it is higher compared to that of a study conducted in Brazil (12,15,18). This might be due to the categorization CD4 count into small intervals in a previous study.

Failure to disclose status puts TB/HIV co-infected patients and their families at higher risk of death. It prevents other at risk family members from being tested and patients who do not disclose their status are highly likely to be lost to follow up, as they cannot explain their ongoing need to visit health institutions (28). In the current study, patients who didn’t disclose their HIV status to anybody were about four times (AHR= 4.03) at higher risk of death than those who disclosed their status. This is in line with the finding of the study done in Jinka, Ethiopia (29). This might be due to patients who didn’t disclose their status would not be comfortable to take their medications appropriately, couldn’t get important family supports and wouldn’t visit health institutions as expected; which might contribute to the unwanted outcome like ART failure and poor survival (30). One of the main reasons for non-disclosure of the status is fear of stigmatization; and this needs a sustainable awareness creation in the community to bring positive attitudes toward TB/HIV co-infected patients and alleviate lay beliefs and misconceptions (8,9).

## Limitation of the study

Selection bias might be introduced; during the exclusion of incomplete registers and subjects with lost cards.

## Conclusions

This study demonstrated short survival time, among TB/HIV co-infected patients during TB treatment, compared to previous findings. A high proportion of deaths occurred within the first two months of anti-TB initiation. In addition, it identified nonuse of CPT, bedridden functional status, nondisclosure of HIV status and CD4 count less than 200 cells/mm^3^ as predictors of time to death among TB/HIV co-infected patients. Close monitoring of bedridden and low CD4 count patients, prompt CPT initiation and encouraging HIV status disclosure are recommended.

## Data Availability

All data are available from corresponding author for any reasonable requests.

https://mail.google.com/wonde1983.ww@gmail.com

## List of abbreviations

AHR: Adjusted Hazard Ratio
AIDS: Acquired Immuno Deficiency Syndrome
AOR: Adjusted Odds Ratio
ART: Anti-Retro Viral Therapy
BMI: Body Mass Index
CD4: Cluster of Differentiation 4
CI: Confidence Interval
CPT: Cotrimoxazole Preventive Therapy
HIV: Human Immuno Deficiency Virus
HR: Hazard Ratio
IRB: Institutional Review Board
OIs: Opportunistic Infections
PLWHA: Peoples Living With HIV/IDS
TB: Tuberculosis
UNAIDS: The joint United Nations Programme on HIV/AIDS
WHO: World Health Organization

## Acknowledgements

We would like to acknowledge the administrative bodies of Mizan Tepi University teaching hospital who allowed us to do this study on their institution and staffs of ART and TB clinics who participated in the collection of data and supervision of its overall processes.

## Author Contributions

**Conceptualization:** Wondimagegn Wondimu

**Formal analysis:** Wondimagegn Wondimu, Teshome Kabeta

**Methodology:** Wondimagegn Wondimu, Teshome Kabeta, Lamessa Dube

**Validation:** Wondimagegn Wondimu, Teshome Kabeta, Lamessa Dube

**Writing original draft:** Wondimagegn Wondimu

**Writing review & editing:** Wondimagegn Wondimu, Teshome Kabeta, Lamessa Dube

